# Socratic AI: An Adaptive Tutor for Clinical Case Based Learning^*^

**DOI:** 10.1101/2025.06.22.25329661

**Authors:** Niloufar Golchini, Evan Passalacqua, Lucy Vaughn, Raja-Elie E. Abdulnour, Travis Zack, Samuel Finlayson

## Abstract

Clinical reasoning is a fundamental skill in medical education that requires intensive faculty resources and deliberate practice. Here, we present the design and implementation of a novel adaptive Socratic tutor powered by large language models (LLMs) that facilitates case-based learning for medical trainees. Our system takes input from any structure of a published clinical case to create interactive and adaptive clinical scenarios where learners engage in realistic patient encounters with real-time feedback on their reasoning process. As a proof of concept, we demonstrate its use with the NEJM clinical pathological case series. This paper describes the architecture, knowledge representation, and educational design principles incorporated into our system, which we are releasing as an open-source tool for medical education. Our work demonstrates how LLMs can promote high-quality precision education in clinical reasoning and provide a structured assessment of the strengths and weaknesses of the learner.

## 1. Introduction

The development of clinical reasoning skills is essential for medical professionals [13], requiring exposure to diverse clinical scenarios and deliberate practice with expert feedback. The path to expertise in clinical reasoning requires approximately 10,000 hours of deliberate practice, yet the average U.S. medical resident completes only 2,500-3,000 hours during their training [1]. This significant gap between required and achieved deliberate practice highlights a critical need for supplementary educational approaches that can provide additional opportunities for structured clinical reasoning development without increasing the already substantial burden on clinical faculty.

Case-based learning (CBL) has been a cornerstone of medical education for over a century, epitomized by the NEJM Clinical Pathological Conference (CPC) since 1924. This methodology, which presents learners with complex clinical scenarios and guides them through the diagnostic process, has proven effective in developing clinical reasoning skills. However, traditional CBL faces several significant limitations in contemporary medical education. Published versions, such as the NEJM CBC, are typically consumed through passive reading, which doesn’t encourage the interaction and engagement that stimulates active learning and long term retention. Faculty availability to facilitate these discussions for guided CBL has always been limited and skewed towards high resource academic settings, resulting in learning disparities in hands-on guidance. Simultaneously, the feedback provided in these sessions, when available, is often inconsistent across instructors, can reflect implicit bias of the educator, and may lack the immediacy and detail necessary for optimal learning. Assessment methodologies in CBL are arduous, inconsistently applied, and subject to significant instructor bias[12], creating challenges in standardizing the evaluation of clinical reasoning competencies.

Finally, the logistics of scheduling and conducting case-based learning sessions limit the frequency with which learners can engage in this valuable form of deliberate practice, creating a significant bottleneck and failing to accommodate the diverse learning rates among students.

Large language models (LLMs) present a unique opportunity to address these gaps by enabling scalable, interactive, and individualized case-based learning. Recent advancements in natural language processing have resulted in AI systems capable of understanding complex medical terminology, maintaining coherent discourse across extended interactions, and providing contextually appropriate responses. Building on these capabilities, we have developed a Socratic AI tutor that guides learners through realistic clinical cases, provides immediate feedback on their reasoning process, and assesses their competencies in real-time.

## 2. Background

Clinical reasoning—the cognitive process physicians use to diagnose and make treatment decisions—relies on integrating biomedical knowledge, pattern recognition, hypothesis testing, and probabilistic thinking[14]. McGaghie et al. have shown that mastery learning with deliberate practice leads to improved clinical performance and better patient outcomes [2]. Structured repetition combined with specific feedback outperforms traditional experience-based training.

Deliberate practice involves four essential elements: motivated learners, well-defined tasks, appropriate difficulty, and informative feedback [3]. In clinical reasoning, this means repeated engagement with diverse clinical scenarios and targeted feedback on the reasoning process itself—not just the final answer.

Unfortunately, current medical education structures often lack sufficient opportunities for this type of deliberate practice. End-of-rotation evaluations typically provide delayed, nonspecific feedback (e.g., ‘great job, read more”), without actionable guidance for improvement. While objective structured clinical examinations (OSCEs) can provide more immediate feedback, they are resource-intensive and limited in scope. Multiple-choice examinations, though efficient to administer, fail to capture the complex thought processes and iterative nature of real clinical reasoning [4].

The Socratic method, another cornerstone of medical teaching, uses guided questioning to develop critical thinking. By encouraging learners to articulate and examine their reasoning, it surfaces thought processes for feedback and supports the development of adaptive expertise. Traditionally, this method is modeled by attending physicians leading case discussions with trainees—an approach valued for its effectiveness but constrained by the need for expert time and individualized interaction.

Trained on extensive textual data, LLMs can engage in context-aware, human-like dialogue, interpret complex medical scenarios, and simulate reasoning patterns. Unlike earlier AI applications that focused on static content delivery or simple assessment, LLMs enable dynamic, adaptive learning experiences [5]. LLMs’ ability to sustain extended dialogue and adjust responses based on learner input makes them well-suited for Socratic-style case-based learning. Romano et al. demonstrated that LLMs can engage in sophisticated medical discourse, interpreting clinical information accurately and generating evidence-based reasoning [5]. This suggests a promising role for LLMs in delivering deliberate practice in clinical reasoning at scale.

## 3. Introduction to the Application

Our system architecture, conceptualized to facilitate adaptive Socratic case-based learning, integrates multiple components that work together. The architecture consists of five key components that manage different aspects of the educational experience (Figure 1).

**Figure 1.**
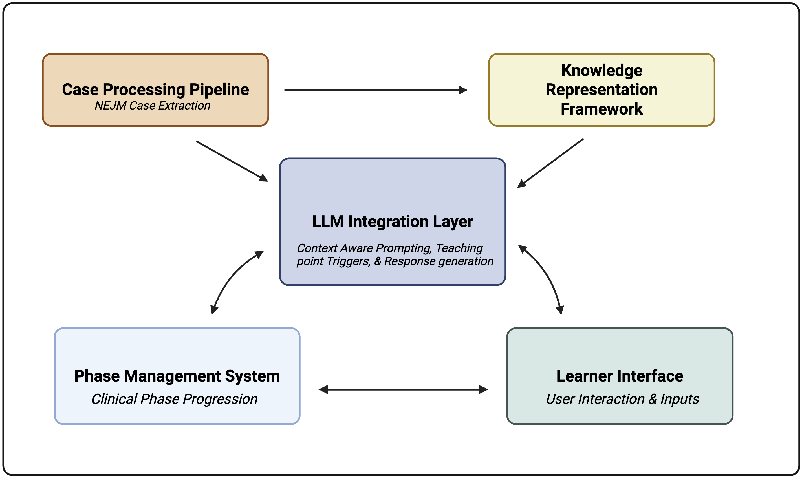
Core Components of the Socratic AI Tutor Architecture

### 3.1 Case Processing Pipeline

The case processing pipeline transforms published clinical cases into structured educational content. To model the iterative nature of clinical reasoning [10], the system breaks down narrative case reports into discrete clinical elements that learners can discover through active inquiry. For example, in a heart failure case, the system separates information about the onset of dyspnea, the severity of orhopnea, and the history of medications into distinct elements that can be revealed individually based on specific questions.

This process preserves the authentic complexity of clinical cases while enabling progressive disclosure of information. When interacting with processed cases, learners experience a realistic simulation of patient interviewing and clinical data gathering—they must ask relevant questions to elicit important information, mirroring real clinical practice where patients don’t automatically volunteer all relevant details.

By preprocessing cases prior to learner engagement, socratic AI identifies and categorizes key teaching opportunities within each case, allowing the system to plan and highlight important clinical concepts, common diagnostic pitfalls, and evidencebased practice points during appropriate moments in the learner’s case exploration.

### 3.2 Knowledge Representation Framework

Our knowledge representation framework organizes clinical case information in a structured format that drives the interactive learning experience. The framework maps patient data that was previously extracted from cases into interconnected clinical elements that can be progressively revealed through learner inquiry.

The framework organizes clinical elements—the discrete pieces of patient information extracted by the processing pipeline—into a semantically rich network (Figure 1). Each element (symptoms, examination findings, laboratory results) is classified according to both medical systems (cardiovascular, respiratory, neurological) and diagnostic significance. For example, in a heart failure case, the framework would represent orthopnea as a respiratory symptom with high specificity for cardiac etiology, while fatigue would be categorized as a constitutional symptom with low diagnostic specificity. This organization creates explicit relationships between clinical findings and potential diagnoses, with weighted connections representing diagnostic relevance.

During interactions, the framework guides the adaptive presentation of information. When a learner asks about a specific symptom, the system doesn’t just provide isolated facts but contextualizes information within its diagnostic framework. If an important connection remains unexplored—such as a learner failing to associate peripheral edema with potential cardiac causes—the system can generate targeted questions that highlight these relationships without explicitly revealing them. Teaching objectives and learner skill level are integrated directly within this clinical network, linking educational goals to specific clinical elements and diagnostic pathways.

When learners discover key clinical findings, the system identifies opportunities to emphasize important concepts. For example, when a learner elicits information about S3 heart sounds, the framework triggers a teaching moment about ventricular dysfunction and volume overload, delivered through Socratic questions rather than didactic statements. This integrated organization allows the system to maintain a comprehensive model of each learner’s progress, tracking not only which information has been discovered but how effectively that information is being integrated into diagnostic reasoning.

### 3.3 Phase Management System

The phase management system guides learners through a structured clinical reasoning process that mirrors authentic practice: hypothesis-driven data acquisition and iterative reasoning. Users progress through five distinct phases—History, Physical Examination, Testing, Management, and Discussion—each representing a key component of clinical decision-making (Figure 2).

**Figure 2.**
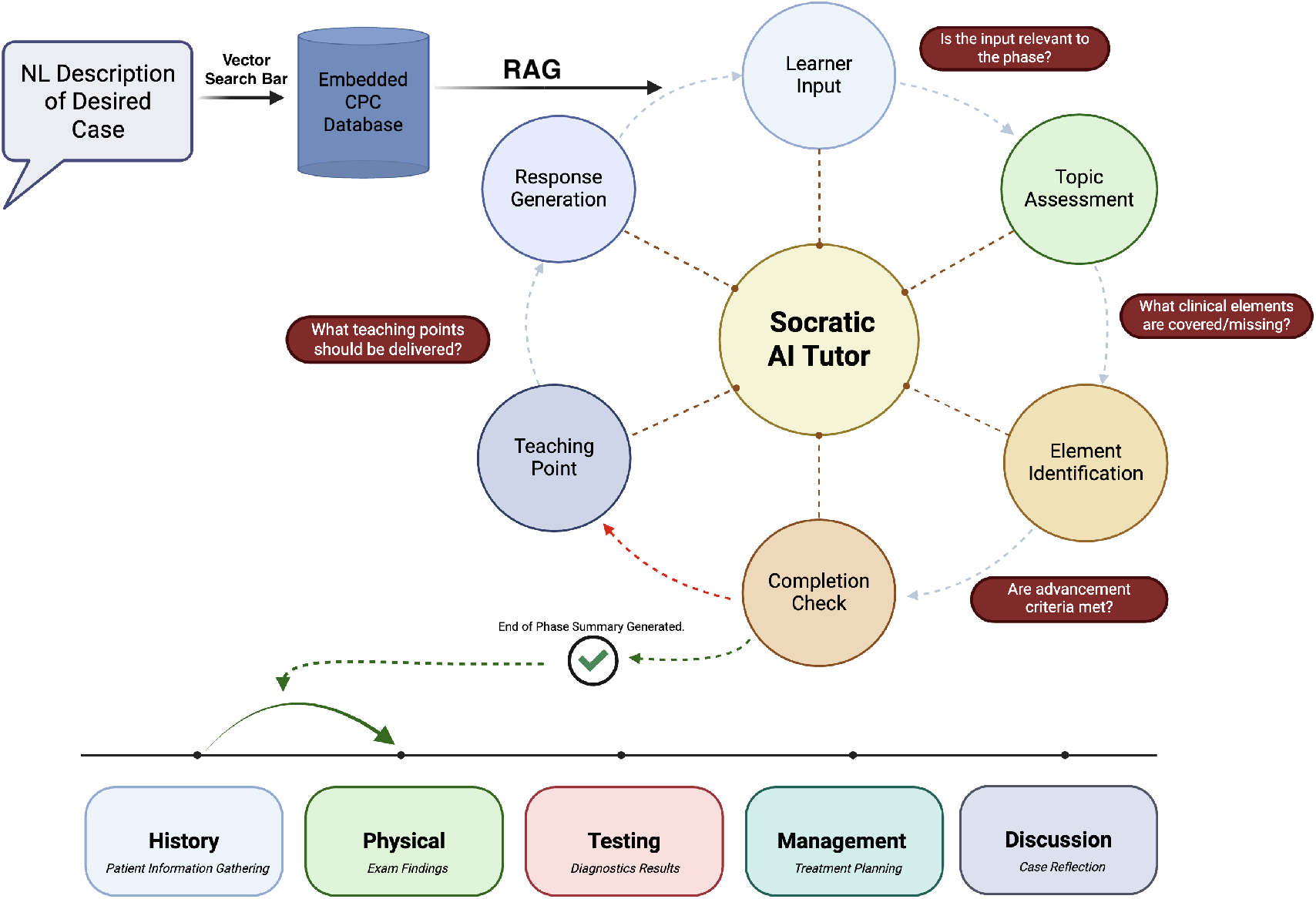
Concept Flow Map of the Feedback Mechanism in the Socratic AI Tutor

**Figure 3.**
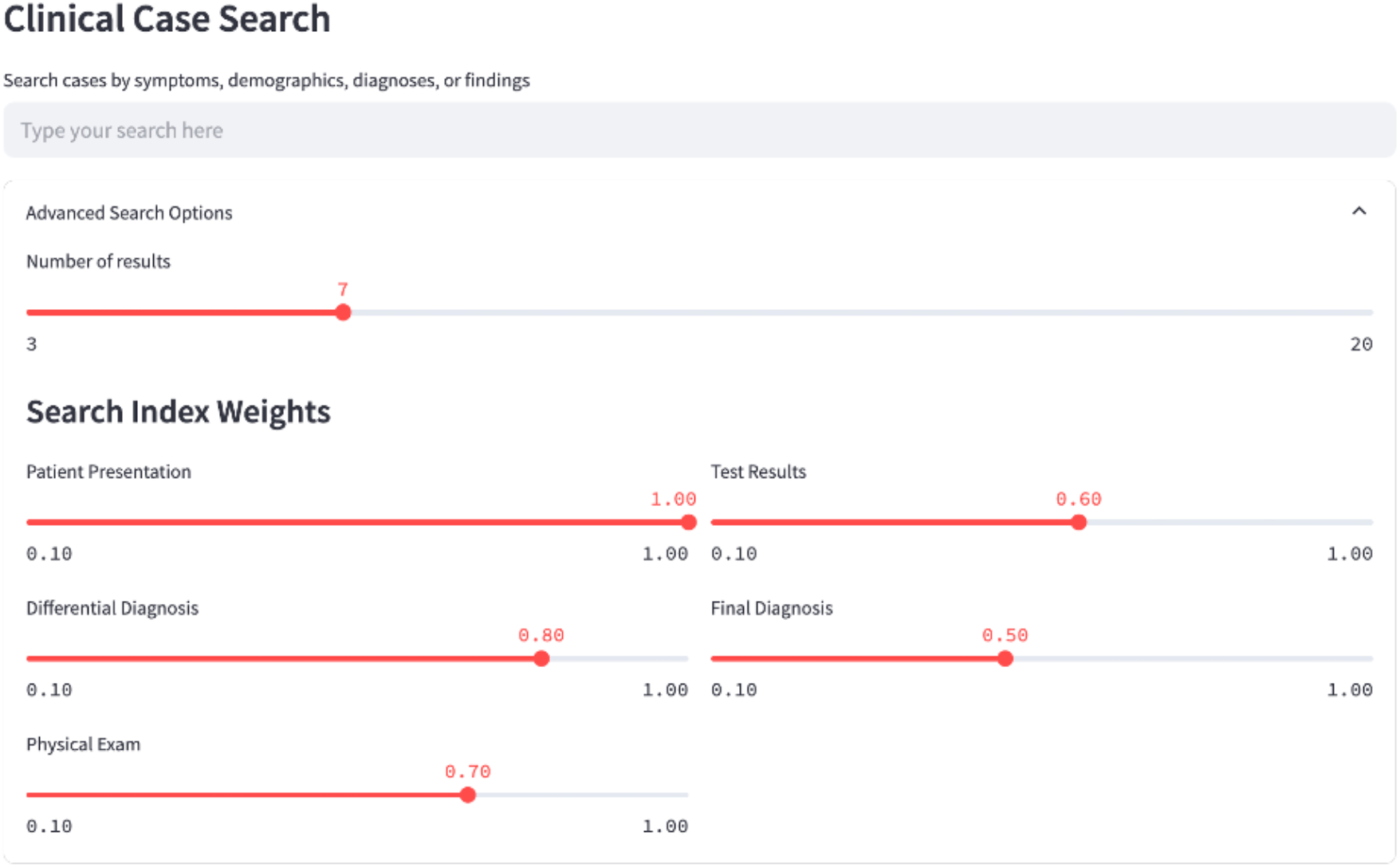
Clinical case vector search bar to find and select a desired case

**Figure 4.**
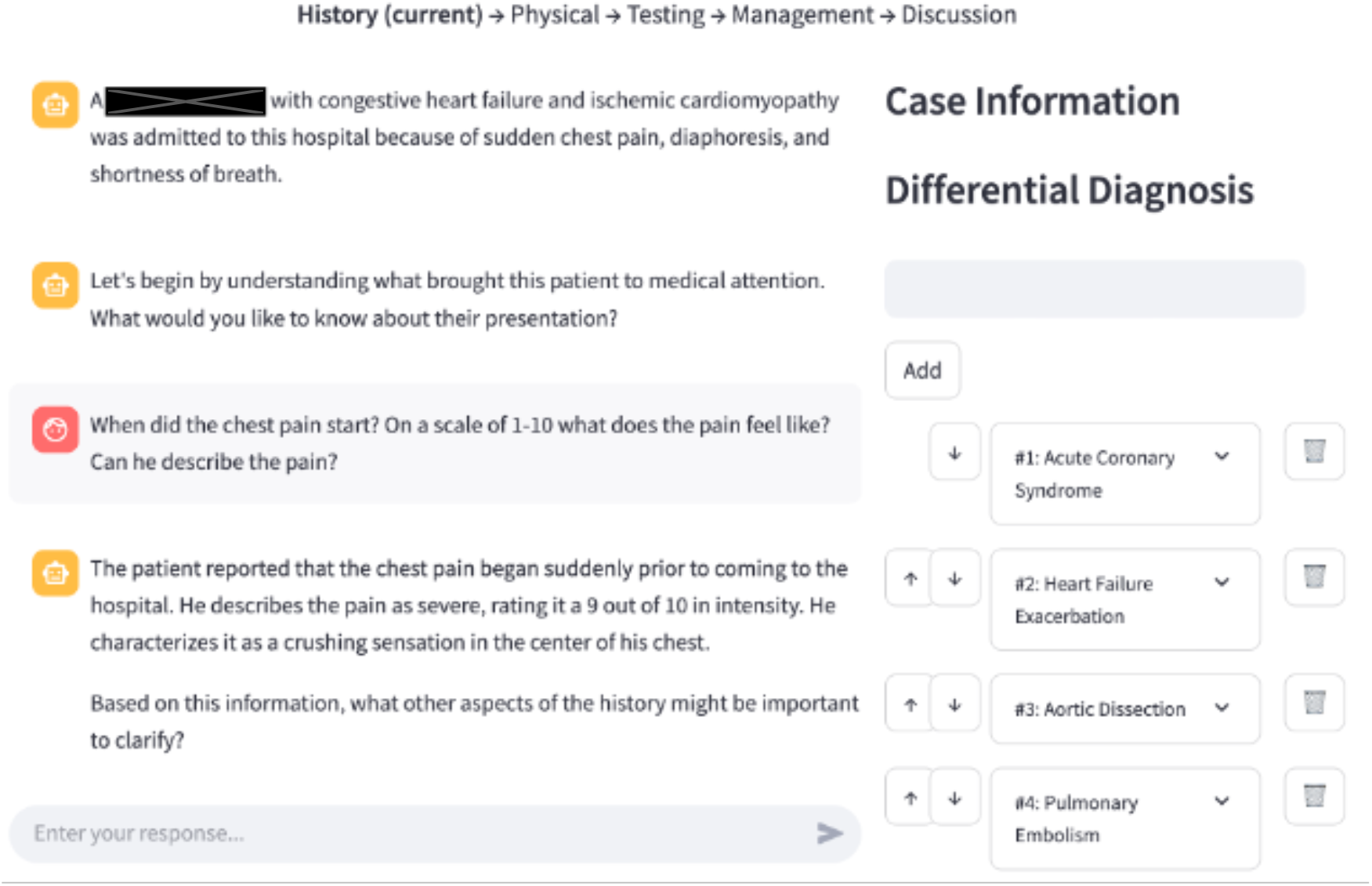
User interface with Socratic AI, including text bar, phase visual indicator and real-time differential diagnosis tracker

**Figure 5.**
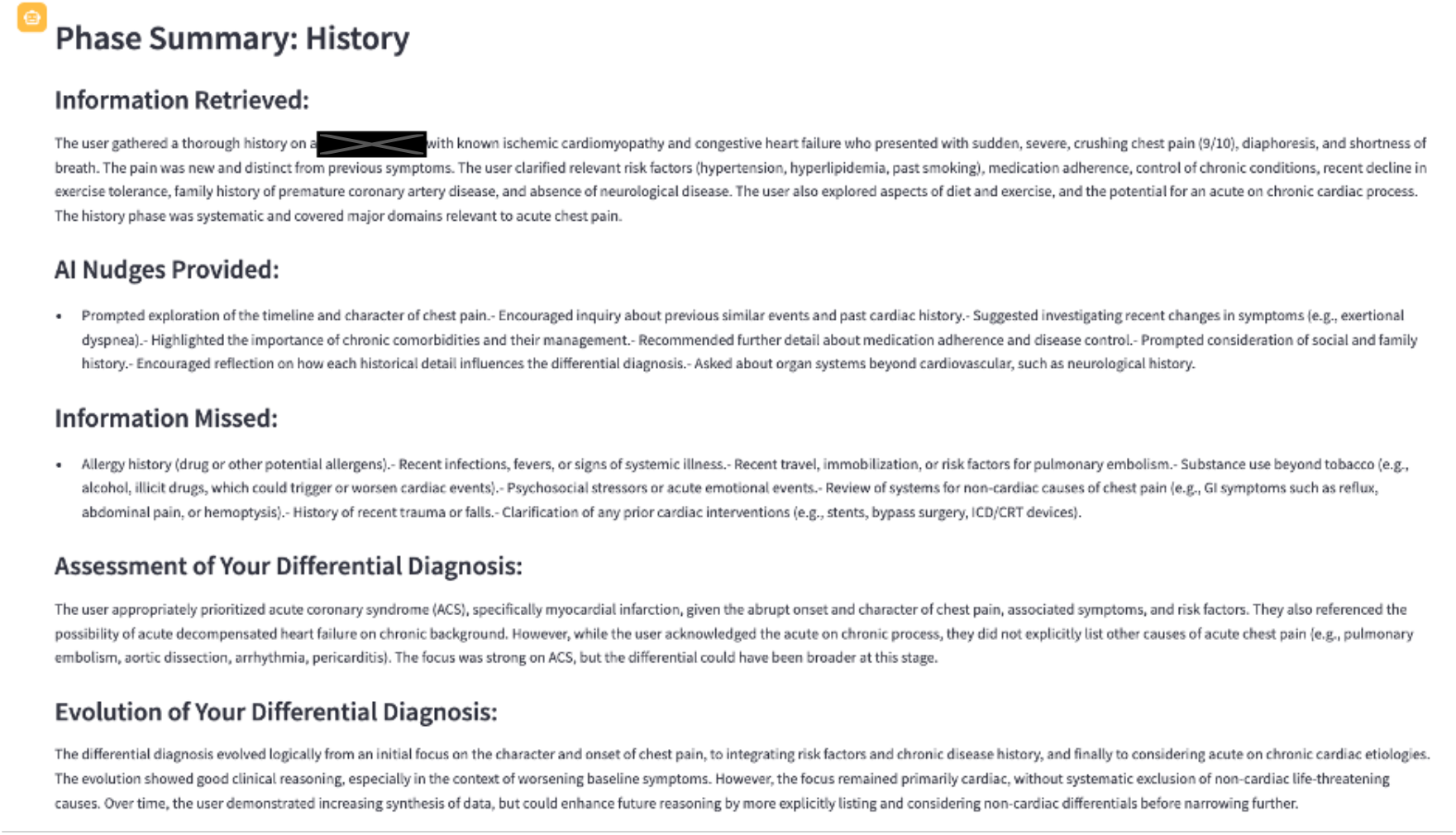
Socratic AI comprehensive end-of-phase summary detailing both educational information and structured feedback in each key dimension

**Figure 6.**
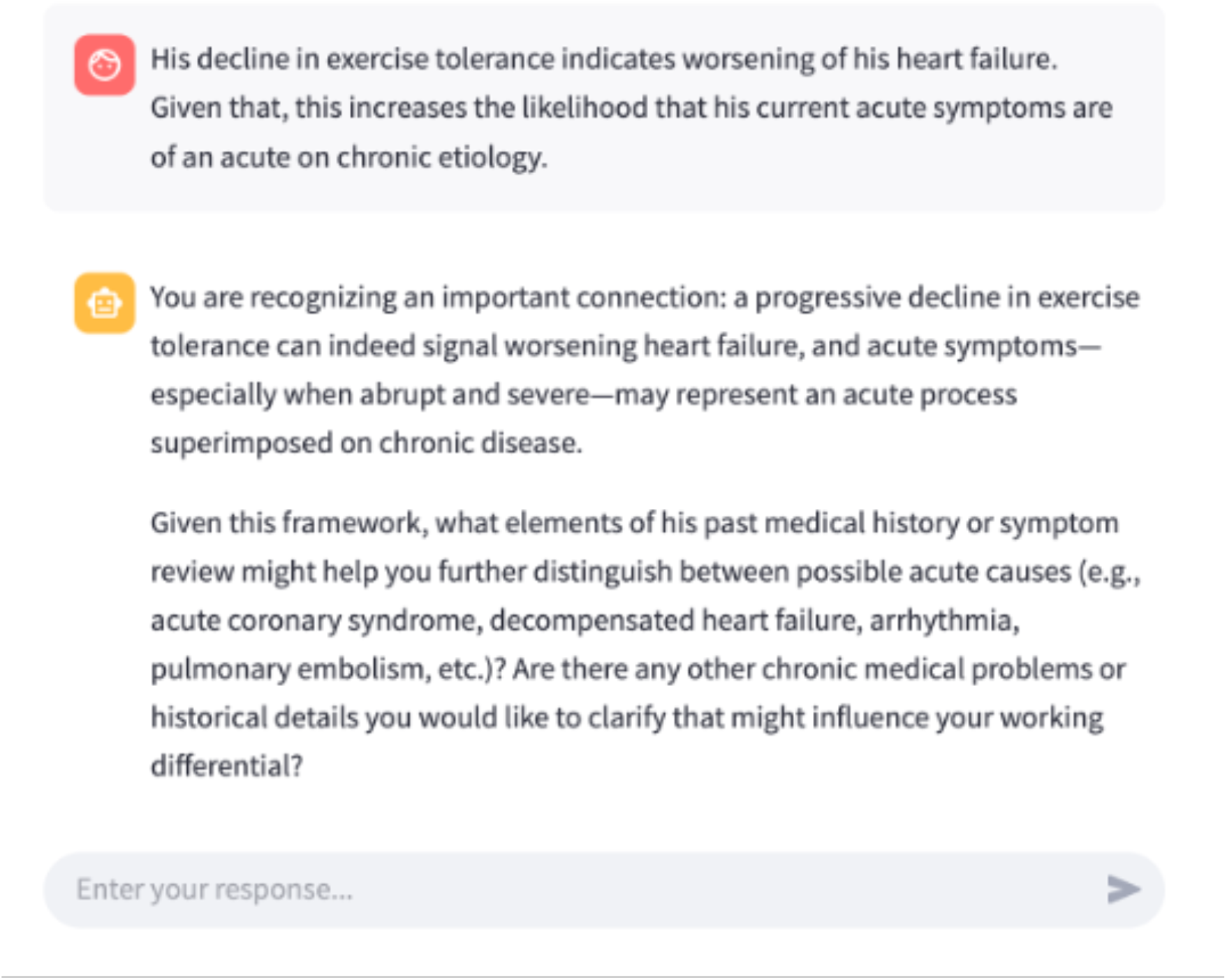
Socratic AI emphasizing important concepts for the user and guiding the user to integrate the concept into their diagnostic reasoning

**Figure 7.**
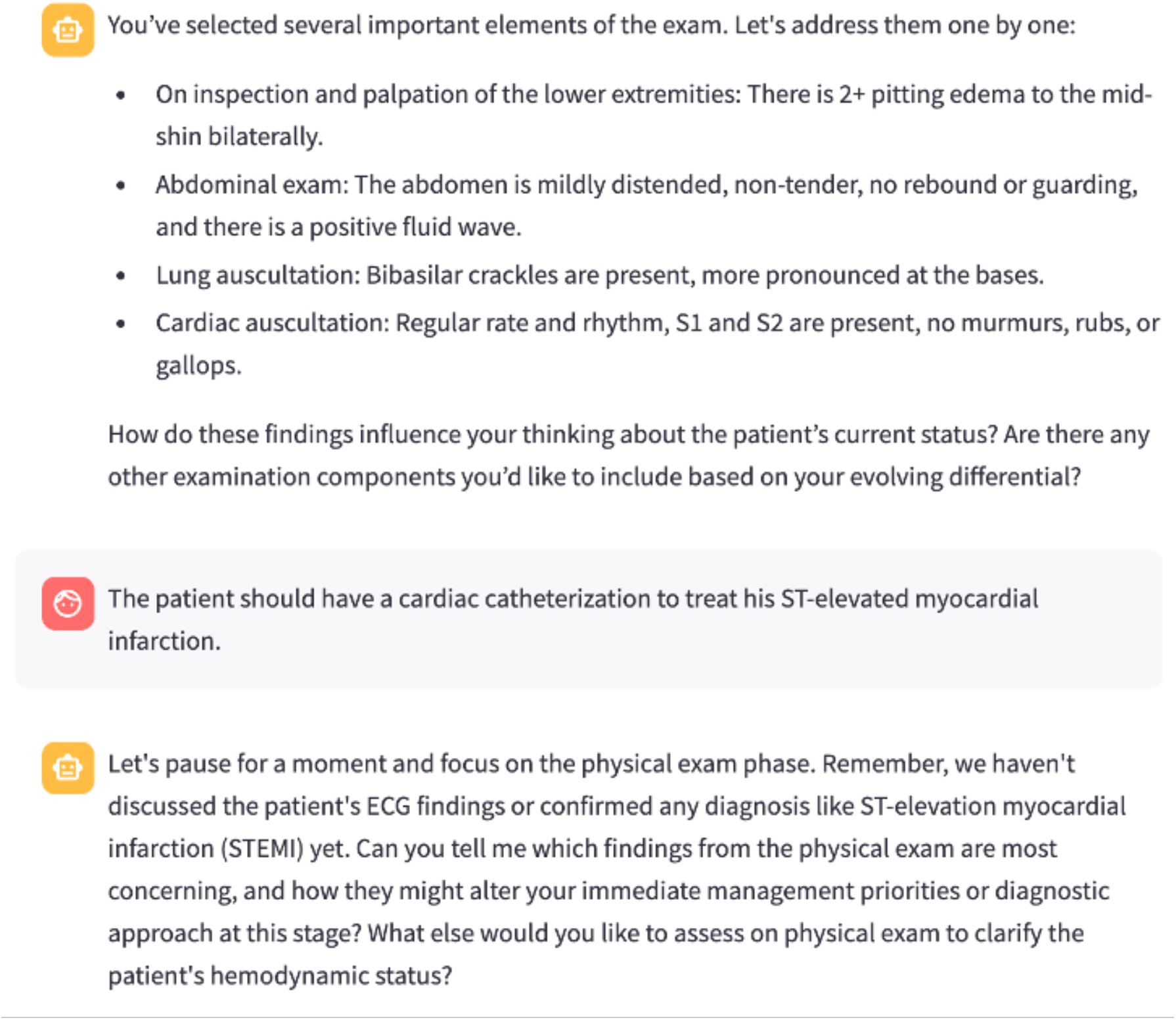
Socratic AI redirects the user when a question diverges from their current phase

**Figure 8.**
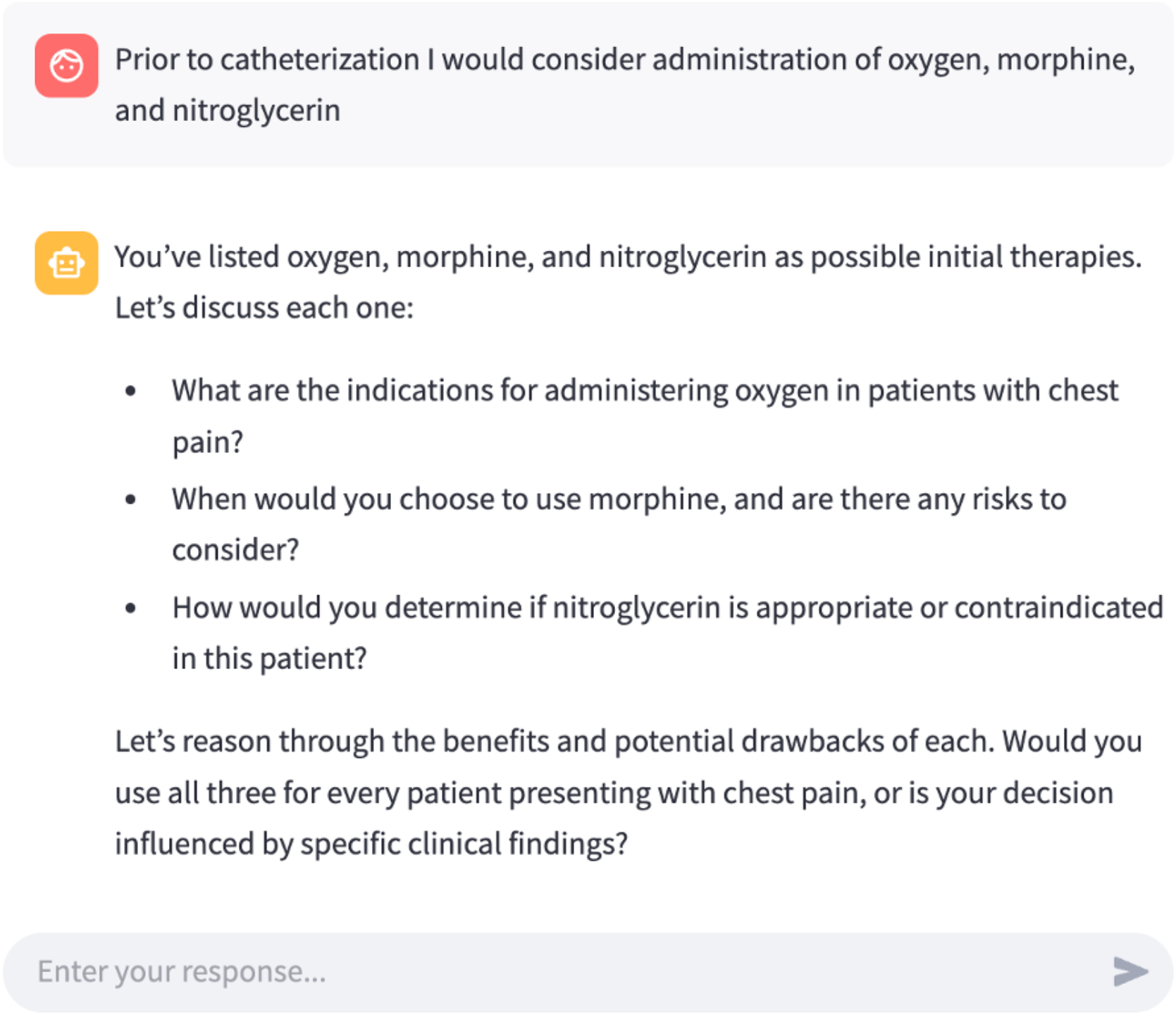
Socratic AI guides the user to develop evidence-based treatment plans and to thoroughly evaluate the benefits and drawbacks of various clinical approaches

When users begin a case, they enter the History phase where they interact with the tutor through natural dialogue. The system responds with appropriate clinical information while implicitly tracking which history elements have been elicited. Users can see their progress through a visual indicator showing which phase they’re currently in, but the specific required elements remain intentionally hidden to preserve authentic exploration.

Socratic AI has an agent that keeps track of the learners progress in eliciting important information for each phase. Once sufficient progress has been made in gathering history, the system enables transition to the Physical Examination phase. Here, users specify which examinations they wish to perform (e.g., “I’d like to check for jugular venous distension” or “Let me examine the lower extremities for edema”), receiving realistic findings that build upon the history they’ve collected. For learners who diverge from the current phase (e.g., asking to order an ECG before completing basic history-taking), the system offers redirection calibrated to their preset guidance level—more direct correction for novice users versus subtle suggestions for advanced learners.

Questions that relate to later phases (e.g., requesting imaging studies during history collection) trigger re-directive responses calibrated to the present guidance level.

In the Testing phase, learners inquire about labs and imaging interventions with the system providing results while subtly guiding them toward appropriate test selection based on their evolving hypotheses. The Management phase challenges learners to develop evidence-based treatment plans, while the Discussion phase promotes reflection on key learning points and alternative diagnostic possibilities.

Throughout this progression, the phase management system continuously evaluates whether learners have gathered essential information before allowing advancement. This approach allows tuning of required comprehensiveness based on level of learner, prevents premature closure and ensures comprehensive exploration while maintaining the feel of a natural clinical encounter.

### 3.4 Adaptive Feedback Mechanisms

The adaptive feedback mechanism responds dynamically to learner interactions, providing contextually appropriate guidance that balances support with challenge.

A key feature of the system is the ability for users to preset their preferred guidance level at the beginning of a case. Learners can select from three guidance tiers:

1. *Novice mode (guidance level high)*: provides more structured support with explicit hints, frequent checks for understanding, and more directive questioning when learners appear to be missing critical information.
2. *Intermediate mode (guidance level medium)*: balances guidance with autonomy, offering occasional suggestions while allowing learners to develop their own approaches to the case.
3. *Advanced mode (guidance level low*): minimizes intervention, challenging experienced learners to independently identify and address knowledge gaps with minimal system assistance.

Within the selected guide level, each input from the learner undergoes a systematic processing workflow that allows contextually appropriate responses (Figure 2). This workflow begins with topic assessment, where the system evaluates whether the learner’s question or comment is relevant to the current clinical phase. For queries appropriate to the current phase (e.g., asking about chest pain characteristics during history-taking), the system proceeds to element identification.

The element identification component analyzes the input of the learner to determine which clinical elements are being addressed and tracks these against the elements required for the current phase. This tracking enables the system to maintain a comprehensive list of explored and unexplored clinical territory. When learners inquire about specific findings, the system not only provides the requested information but also updates its internal model of their progress. Following element identification, the completion check component evaluates whether learners have gathered sufficient information to satisfy phase advancement criteria.

The teaching point component identifies opportunities for educational intervention based on learner interactions. When learners discover findings with significant teaching value or demonstrate reasoning patterns that warrant guidance, this component triggers appropriate Socratic questions or explanations calibrated to the preset guidance level. The system continuously identifies knowledge gaps by tracking unexplored critical elements. When important information remains undiscovered, it generates contextual prompts with varying levels of specificity based on the preset guidance mode. In novice mode, the system might ask, “Have you considered exploring the patient’s cardiac risk factors?” while in advanced mode, it might simply note, “You seem to be developing a good understanding of the respiratory symptoms—are there other systems you’d like to explore?”

A last component of the adaptive support mechanism is the comprehensive end-of-phase summary provided when learners complete each clinical phase. Upon gathering sufficient information to satisfy phase completion criteria, the system notifies learners that they’ve covered the essential elements and offers to generate a phase summary. These summaries serve both educational and assessment functions, providing structured feedback across five key dimensions:

#### Retrieved Information

A comprehensive overview of the clinical data successfully gathered.

#### AI Nudges Provided

A reflection on the guidance offered during the interaction, helping learners recognize where they needed additional support.

#### Missed Information

Identification of critical elements that were overlooked or incompletely explored.

#### Differential Diagnosis Assessment

Evaluation of the quality and evolution of the learner’s diagnostic hypotheses, with feedback on both the content and the reasoning process.

#### Differential Diagnosis Evolution

Analysis of how the learner’s diagnostic thinking developed throughout the phase.

These end-of-phase summaries create progressive documentation of the learner’s clinical reasoning development throughout the case. Users can review these summaries in the information panel at any time, using them to reflect on their progress and approach as they advance through subsequent phases.

## 4. Implementation

### 4.1 LLM Integration and Prompting Strategy

We implemented our system using OpenAI’s GPT-4o. Unlike traditional chatbots that simply respond to queries, our implementation guides the LLM to function as an educational facilitator. The system uses a base instruction set that establishes the LLM’s role as a clinical educator who asks guiding questions rather than providing immediate answers. When a learner interacts with the system, their input is processed alongside contextual information about the current phase, previously covered elements, and pending teaching points. This contextualized prompting enables the LLM to maintain awareness of the educational progression throughout the case.

Phase-specific prompts modify the LLM’s behavior based on the current stage of the clinical encounter. During history-taking, the system adopts a patient-like response style, answering questions about symptoms and history while occasionally demonstrating realistic patient behaviors like uncertainty about medical details. In the testing phase, the system shifts to a more analytical mode, presenting laboratory and imaging results with appropriate clinical interpretation guidance.

Our implementation includes specialized prompting to manage uncertainty appropriately. The LLM is guided to express appropriate confidence levels based on the strength of clinical evidence, modeling the nuanced judgment that characterizes expert clinicians when facing ambiguous presentations.

### 4.2 User Interface

A distinctive feature of the interface is the real-time differential diagnosis tracking system, which provides learners with visual representation of their evolving diagnostic thinking. This component allows learners to add, remove, or reorder diagnostic possibilities as they gather additional information, providing explicit externalization of their reasoning process. The tracking system integrates with the educational framework to provide subtle guidance on diagnostic possibilities that warrant consideration based on the information gathered.

## 5. Discussion

### 5.1 Educational Implications

Our Socratic AI tutor represents a significant advancement in clinical reasoning education, addressing several persistent challenges that have limited the effectiveness of traditional approaches. The implications of this work extend beyond the specific implementation to broader considerations regarding the role of AI in medical education and the future of clinical reasoning instruction.

The system directly addresses the accessibility challenge that has traditionally limited high-quality clinical reasoning instruction. By providing ondemand access to interactive clinical scenarios, the system democratizes access to high-quality educational experiences that were previously available only to learners at institutions with sufficient expert faculty resources. This expanded accessibility has particularly significant implications for educational equity, potentially reducing disparities in clinical reasoning education between resource-rich and resource-constrained institutions.

Our system creates unprecedented opportunities for deliberate practice outside traditional clinical settings. Learners can engage with diverse clinical scenarios at their convenience, receiving immediate feedback on their reasoning process without requiring faculty presence. This flexibility enables more frequent engagement with clinical reasoning exercises, potentially accelerating skill development through increased practice volume. The system’s ability to present multiple variations of similar clinical scenarios is particularly valuable for developing adaptive expertise, as described by Mylopoulos et al. [6], helping learners recognize the same condition across different presentations.

Existing screen based virtual patient simulators provide learners with on-demand access to interactive clinical scenarios and expert-level feedback, with the potential to significantly enhance clinical reasoning development [11]. Our Socratic AI tutor builds upon this foundation by incorporating adaptive questioning and real-time feedback that simulates the guidance of an expert preceptor. This enhancement potentially bridges the gap between static case presentations and authentic clinical learning, creating a more engaging and effective educational experience.

### 5.2 Limitations and Future Directions

While our Socratic AI tutor demonstrates significant promise for enhancing clinical reasoning education, we acknowledge several important limitations that must inform interpretation of our results and guide future development.

The system’s inability to fully replicate the nuances of human expert guidance represents a fundamental limitation. Despite sophisticated natural language capabilities, the LLM cannot match the emotional intelligence, experiential wisdom, and intuitive understanding that characterize expert human educators. The system cannot detect subtle non-verbal cues from learners, such as confusion or frustration, that might prompt a human educator to adjust their approach. Most importantly, it does not substitute the mentorship and human connection between experienced physician and trainee that case based learning can facilitate. This limitation underscores the complementary rather than replacement role of AI in clinical education.

The potential reinforcement of biases present in training data represents a significant concern requiring ongoing attention. The system may inadvertently perpetuate biases present in the medical literature and case repositories from which it draws. These biases might manifest in various forms, including over- or under-representation of certain demographic groups in particular clinical scenarios or subtle differences in language when describing similar presentations in different populations.

The system’s dependency on the quality of underlying case materials represents both a limitation and an area for future development. While the NEJM case repository provides high-quality source material, it necessarily reflects the perspectives, biases, and limitations of its contributors. The cases may not represent the full diversity of clinical presentations or practice contexts, potentially limiting the breadth of educational exposure. Expanding the system to incorporate more diverse case sources while maintaining quality standards represents an important future direction.

## 6. Conclusion

While acknowledging important limitations regarding the replication of human expertise, potential biases, assessment of non-verbal skills, and dependence on case quality, we believe that the system represents a valuable addition to the clinical education toolkit rather than a replacement for human teaching. The complementary role of AI-enabled learning systems alongside traditional approaches offers the potential to enhance overall educational effectiveness while addressing persistent challenges in clinical reasoning education. This work establishes a foundation for future AI-assisted medical education tools and contributes to our understanding of how technology can enhance the development of complex cognitive skills in healthcare professionals.

## Data Availability

All data produced in the present study are available upon reasonable request to the authors, and will be made open-source upon peer-reviewed publication.

## 7. Acknowledgments

This work was supported by the NAIRR Pilot Program under award number NAIRR240071, “An adaptive large language model for Socratic case-based learning in medical education.” We gratefully acknowledge the computational resources provided through this award, including support from Microsoft Azure and TACC Lonestar6, which enabled our use of large language models in this research.

## 8. Appendix

The Socratic AI Agentic system uses OpenAI’s gpt-4o model to handle all generative tasks, including clinical tutoring and summarization, with temperature adjusted based on use case (e.g., 0.2 for search summaries, 0.7 for conversational prompts). Embeddings are computed using text-embedding-3-large.

## 8.1 Retrieval-Augmented Case Search

To support users in finding clinically relevant examples from a growing database of cases, we implemented a retrieval-augmented generation (RAG) search bar within the Clinical Case Tutor. This feature allows users to input natural language queries and receive ranked matches based on semantic similarity to indexed patient data, promoting exploration and pattern recognition across cases.

The system uses OpenAI’s embedding models to generate high-dimensional vector representations of several aspects of each case: patient presentation (PoC), differential diagnosis (DDx), physical examination (PD), test results (TD), and final diagnosis (FD). Each aspect is embedded independently, enabling granular semantic retrieval based on the most relevant clinical dimension. For example, a user interested in comparing differential diagnoses may assign greater search weight to DDx vectors while de-emphasizing final outcomes.

Before embedding, each user query is normalized using an LLM step that standardizes clinical language. This includes converting lay terms and abbreviations into medically precise language (e.g., “heart attack” becomes “myocardial infarction”), improving alignment between the query and embedded case representations.

These vectors are indexed using FAISS (Facebook AI Similarity Search), which allows an efficient approximate nearest neighbor search across cases. At query time, the normalized query is embedded using the same model. The resulting vector is searched against each FAISS index, retrieving top matches by cosine similarity.

Results are scored using a weighted relevance formula:

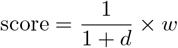

where *d* is the cosine distance and *w* is the preassigned weight for that clinical dimension. Default weights prioritize patient presentation (*w* = 1.0) and differential diagnoses (*w* = 0.8), with lower contributions from physical, test, and final diagnosis dimensions (*w* = 0.7, 0.6, and 0.5, respectively). These weights are user-configurable through the search interface, allowing for contextual emphasis based on the user’s learning objective.

The system aggregates results across all dimensions and returns the top-*k* scoring cases. To assist interpretation, the application generates a brief LLM-authored summary contextualizing the retrieved cases, highlighting shared features and potential patterns of clinical significance. This summary is produced at low temperature to ensure factual consistency.

